# Safety and Immunogenicity of an Inactivated Recombinant Newcastle Disease Virus Vaccine Expressing SARS-CoV-2 Spike: A Randomised, Comparator-Controlled, Phase 2 Trial

**DOI:** 10.1101/2023.11.30.23299208

**Authors:** Vu Dinh Thiem, Dang Duc Anh, Vu Hai Ha, Nguyen Van Thom, Tran Cong Thang, Jose Mateus, Juan Manuel Carreño, Rama Raghunandan, Nguyen Mai Huong, Laina D Mercer, Jorge Flores, E Alexandar Escarrega, Ariel Raskin, Duong Huu Thai, Le Van Be, Alessandro Sette, Bruce L Innis, Florian Krammer, Daniela Weiskopf

## Abstract

**Background:** Production of affordable coronavirus disease 2019 (COVID-19) vaccines in low- and lower-middle-income countries is needed. NDV-HXP-S is an inactivated egg-based recombinant Newcastle disease virus vaccine expressing the spike protein of severe acute respiratory syndrome coronavirus 2 (SARS-CoV-2). A public sector manufacturer in Vietnam assessed the immunogenicity of NDV-HXP-S (COVIVAC) relative to an authorized vaccine.

**Methods:** This phase 2 stage of a randomised, observer-blind, controlled, phase 1/2 trial was conducted at three community health centers in Thai Binh Province, Vietnam. Healthy males and non-pregnant females, 18 years of age and older, were eligible. Participants were randomised by age (18-59, ≥60 years) to receive one of three treatments by intramuscular injection twice, 28 days apart: COVIVAC at 3 µg or 6 µg, or AstraZeneca COVID-19 vaccine VAXZEVRIA. Participants and personnel assessing outcomes were masked to treatment. The main outcome was the induction of 50% neutralising antibody titers against vaccine-homologous pseudotyped virus 14 days (day 43) and 6 months (day 197) after the second vaccination by age group. The primary immunogenicity and safety analyses included all participants who received one dose of the vaccine. ClinicalTrials.gov NCT05940194.

**Findings:** During August 10-23, 2021, 737 individuals were screened, and 374 were randomised (124-125 per group); all received dose one, and three missed dose two. On day 43, the geometric mean fold rise of 50% neutralising antibody titers for subjects age 18-59 years was 31·20 (COVIVAC 3 μg N=82, 95% CI 25·14-38·74), 35·80 (COVIVAC 6 μg; N=83, 95% CI 29·03-44·15), 18·85 (VAXZEVRIA; N=82, 95% CI 15·10-23·54), and for subjects age ≥60 years was 37·27 (COVIVAC 3 μg; N=42, 95% CI 27·43-50·63), 50·10 (COVIVAC 6 μg; N=40, 95% CI 35·46-70·76), 16·11 (VAXZEVRIA; N=40, 95% CI 11·73-22·13). Among subjects seronegative for anti-S IgG at baseline, the day 43 geometric mean titer ratio of neutralising antibody (COVIVC 6 μg/VAXZEVRIA) was 1·77 (95% CI 1·30-2·40) for subjects age 18-59 years and 3·24 (95% CI 1·98-5·32) for subjects age ≥60 years. On day 197, the age-specific ratios were 1·11 (95% CI 0·51-2·43) and 2·32 (0·69-7·85). Vaccines were well tolerated; reactogenicity was predominantly mild and transient. The percentage of subjects with unsolicited adverse events (AEs) during 28 days after vaccinations was similar among treatments (COVIVAC 3 μg 29·0%, COVIVAC 6 μg 23·2%, VAXZEVRIA 31·2%); no vaccine-related AE was reported.

**Interpretation:** Considering that induction of neutralising antibodies against SARS-CoV-2 has been correlated with the efficacy of COVID-19 vaccines, including VAXZEVRIA, our results suggest that vaccination with COVIVAC may afford clinical benefit matching or exceeding that of the VAXZEVRIA vaccine.

**Funding:** Vietnam’s Institute of Vaccines and Medical Biologicals (including support from Vietnam’s national COVID-19 vaccine fund and a charitable contribution from the Thien Tam fund of Vin group), Coalition for Epidemic Preparedness Innovations, a charitable contribution from Bayer AG, US National Institutes of Health.

## Introduction

The coronavirus disease 2019 (COVID-19) pandemic has resulted in millions of deaths, burdened healthcare systems globally, and exposed vaccine access inequities worldwide. A systematic study to assess the impact of delayed supply of COVID-19 vaccines indicated that only 25% of the population in low- and lower-middle-income countries received at least one dose of vaccine as of October 2022.^1^ Ensuring an adequate supply of COVID-19 vaccines for low- and lower-middle-income countries (LMICS), which constitute 85% of the global population, is essential.

As of March 2023, Vietnam’s Ministry of Health recorded 11,525,408 COVID-19 cases, ranking 13^th^ in amount of cases among 230 countries and territories worldwide.^2^ Although imported vaccines and infection-induced immunity have reduced the risk of disease, the threat from new viral variants and the potential need for vaccinating elderly adults and other at-risk individuals annually highlight the value to Vietnam of access to domestically produced COVID-19 vaccines as a sustainable asset.

The rapid rollout of COVID-19 vaccines saved millions of lives globally.^3^ By inducing potent severe acute respiratory syndrome coronavirus 2 (SARS-CoV-2) antibodies, COVID-19 vaccines reduce the risk of severe disease, with the level of antibodies induced correlated with vaccine efficacy.^4-5^ However, the emergence of Omicron sub-lineage variants with increased transmissibility and escape from pre-existing neutralising antibodies emphasizes the importance of confirming that new COVID-19 vaccine candidates also induce cellular immunity.^6^

PATH and the Icahn School of Medicine at Mount Sinai collaborated with Vietnam’s Institute of Vaccines and Medical Biologicals (IVAC), a manufacturer of egg-based inactivated influenza vaccines, to develop an egg-based inactivated Newcastle disease virus vaccine expressing a six-proline prefusion-stabilized SARS-CoV-2 spike (NDV-HXP-S COVID-19 vaccine, also known as COVIVAC).^7^ In a phase 1 trial (NCT04830800), COVIVAC administered twice 28 days apart had an acceptable safety and immunogenicity profile in healthy adults 18-59.^8^ For the next stage of clinical development, IVAC sponsored a phase 2 trial in which the safety and immunogenicity of COVIVAC at two dosage levels, in adults with stable health including individuals ≥60 years of age, was contrasted with AstraZeneca’s adenovirus vectored COVID-19 vaccine (VAXZEVRIA)^9^ then the authorized pandemic vaccine most commonly administered in Vietnam. The study aimed to demonstrate that COVIVAC induced a superior neutralising antibody response to vaccine-homologous SARS-CoV-2 relative to VAXZEVRIA. The study also aimed to explore the activation of SARS-CoV-2 spike-specific T cells by COVIVAC versus VAXZEVRIA. This report provides the results of that clinical trial, including the induction of virus neutralising antibodies against pseudotyped and wild-type (live virus) vaccine-homologous SARS-CoV-2 and virus-specific T-cell activation

## Methods

### Study design and participants

This phase 2 stage (NCT05940194) of a randomised, observer-blind, controlled, phase 1/2 trial was conducted at three community health centers within the Vu Thu District Health Center catchment, Thai Binh Province, Vietnam. Investigators from Vietnam’s National Institute of Hygiene and Epidemiology (NIHE) collaborated with staff of the community health centers, district health center, district hospital, and the Provincial Center for Disease Control to perform the study. Participants were recruited following community outreach. Males and non-pregnant females (sex or gender was self-reported) with stable health, 18 years of age and older, with body mass index 17 to 40 kg/m^2^, with no history of confirmed COVID-19 or infection with human immunodeficiency virus, were eligible to participate. A negative urinary pregnancy test was required of women with reproductive capacity before administering each study vaccine dose. Complete eligibility criteria are described in the trial protocol provided in the supplementary material.

Written informed consent was obtained from all participants. The trial complied with the Declaration of Helsinki and Good Clinical Practice. This study was jointly approved by the Institutional Review Board of the Vietnam National Institute of Hygiene and Epidemiology and the Independent Ethics Committee of the Vietnam Ministry of Health ref no. 1407/QD-BYT.

### Randomisation and masking

Subjects (N=374) were randomly allocated to one of three equal groups (COVIVAC 3 µg, COVIVAC 6 µg, or the comparator VAXZEVRIA) using a computer-generated randomisation sequence prepared by an unblinded statistician. Randomisation was age-stratified, with approximately one-third of subjects aged ≥60 years. An unmasked pharmacist dispensed each treatment according to the randomisation sequence to an unmasked vaccinator. All participants and study personnel, besides the unmasked pharmacy team and vaccinators, were masked for treatment.

### Procedures

The recombinant NDV-HXP-S vaccine (COVIVAC) was manufactured according to current Good Manufacturing Practice (GMP) by IVAC in their Influenza Vaccine Plant (Nha Trang, Vietnam), as previously described.^8^ The adenovirus vectored vaccine from AstraZeneca (ChAdOx1; VAXZEVRIA), used as a comparator vaccine, was sourced from the Ministry of Health. Unmasked vaccinators administered study treatments by intramuscular injection of 0·5 mL on study days 1 and 29. Subjects were observed in the clinic for 30 min after each vaccination. Blood samples were drawn for immunogenicity endpoints before vaccination on days 1 (first dose), 43 (14 days post dose two), and 197 (6 months post dose two). Subjects randomly allocated to a cell-mediated immunity subset (N=12 per treatment group) had additional blood collected on days 1 and 43 to isolate peripheral blood mononuclear cells (PBMCs); these were stored in liquid nitrogen until analysed. Solicited injection site reactions (pain/tenderness, swelling/induration, erythema) and systemic symptoms (headache, fatigue, malaise, myalgia, arthralgia, nausea/vomiting, and fever defined as oral temperature ≥38°C) were recorded by subjects in a diary card for seven days post-vaccination that included intensity, which the investigators then reported. Subjects also recorded unsolicited adverse events (AEs) for 28 days after each vaccine dose and reported them at scheduled clinic visits, whereupon the investigator included these in the study database after interviewing the subjects, grading them for intensity as previously described,^8^ assessing them for causality, and categorizing them as severe or not. Severe AEs were collected for the duration of the study. A Data Safety Monitoring Board (DSMB) monitored unblinded safety data.

We measured anti-SARS-CoV-2 spike IgG using a validated indirect enzyme-linked immunosorbent assay (ELISA) at Nexelis (Laval, Canada), as described.^8^ Concentrations were transformed to binding antibody units per mL (BAU/mL), based on the World Health Organization (WHO) International Standard for anti-SARS-CoV-2 immunoglobulin using a conversion factor determined during assay validation (1/7·9815). The assay’s cut-off and lower limit of quantitation (LLOQ) were 6·3 BAU/mL.

We measured serum neutralising activity against the Wuhan-Hu-1 strain of SARS-CoV-2 in a validated pseudotyped virus neutralization assay (PNA)^8^ that assessed particle entry inhibition.^10^ The neutralising titer of a serum sample was calculated as the reciprocal serum dilution corresponding to the 50% neutralisation antibody titer (NT50) for that sample; the NT50 titers may be transformed to international units per mL (IU/mL), based on the WHO international standard for anti-SARS-CoV-2 immunoglobulin, using a conversion factor determined during assay validation (1/1·872). The assay’s cut-off and lower limit of quantitation (LLOQ) were 5·3 IU/mL (10 as the NT50 titer value) and 5·9 IU/mL, respectively.

We also measured live virus neutralising activity as a 50% inhibitory dilution (ID50) against a wild-type SARS-CoV-2 isolate (USA-WA1/2020, catalog number NR-52281; BEI Resources) using an assay performed in a biosafety level 3 facility as previously described.^11^ Briefly, Vero.E6 cells (20,000 cells/100 μL per well) were seeded onto sterile 96-well cell culture plates a day prior to the neutralisation assay. Sera were serially diluted in minimal essential medium (MEM; Life Technologies) at a 1:10 starting dilution. One thousand (1,000) median tissue culture infectious doses (TCID50s) of the virus were incubated with diluted sera for 1 hour inside a biosafety cabinet. Media from confluent cell monolayers (90%) was removed, and 120 μL of the virus-serum dilutions were added to the cells for 1 h at 37°C. The mixture was removed and 100 μL of each corresponding serum dilution was added per well. Additionally, 100 μL of MEM was added to every well. Remdesivir at 10 μM was used as control. Plates were incubated at 37°C for 48 hours, media was removed, and cells were fixed with 150 μL of 10% formaldehyde (Polysciences) per well. After fixation, cells were permeabilized and stained using the 1C7C7 mAb.^11^ The live virus neutralisation assay (LVNA) cutoff (ID50) was 1:10.

To assess the breadths of the adaptive immune response, we measured vaccine-induced spike-specific T cells in PBMC samples utilizing a T cell receptor (TCR) dependent activation induced markers (AIM) assay.^12-13^ AIM assays have been comprehensively used to compare COVID-19 vaccine-induced T cell responses.^14-15^ This assay measures antigen specific T cells based on upregulation of activation markers, irrespective of cytokines.^16^ Antigen-specific CD4+ and CD8+ T cells were measured as a percentage of AIM+ T cells+ as described before.^14,16-17^ Briefly, PBMC were thawed and plated in 96-wells U-bottom plates at 1×10^6^ PBMC per well, then blocked at 37°C for 15 min with 0·5 µg/ml anti-CD40 mAb (Miltenyi Biotec), and fluorescently labeled with chemokine receptor antibodies (anti-CCR6, CXCR5, CXCR3, and CCR7) (see Supplement Table 1 for list of antibodies used). Cells were incubated at 37°C for 24 hrs with a spike-specific peptide mega pool (MP; 1 µg/ml); controls were dimethyl sulfoxide (DMSO, an equimolar amount) and phytohaemagglutinin PHA (2·5 µg/ml). The mega pool (MP) approach, previously described, enables simultaneous testing of a large number of epitopes, facilitating the characterization of T-cell responses to infectious diseases.^13-14^ We stimulated the PBMCs *ex vivo* to evaluate the antigen-specific T cell response against SARS-CoV-2. The spike MP has 253 overlapping peptides spanning the entire sequence of the spike protein.^18^ SARS-CoV-2 spike-specific circulating CD4+ T cells and spike-specific circulating CD8+ T cells were measured by surface co-expression of OX40+CD137+ and CD69+CD137+, respectively. SARS-CoV-2 spike-specific circulating follicular helper T (cTFH) cells were measured as CXCR5+OX40+surface CD40L+ and quantified as a percentage of CD4+ T cells after stimulation with spike MP. The samples were acquired on a Cytek Aurora (Cytek Biosciences). The gating strategy is shown in Supplement Figure 1.

### Outcomes

The primary outcomes were safety and induction of neutralising antibodies by COVIVAC, comparing 3 µg to 6µg and each COVIVAC group to the VAXZEVRIA group. The safety of each treatment was evaluated as the number and severity of solicited injection site and systemic AEs during 7 days after vaccination. Number, severity, and relatedness of unsolicited (spontaneously reported) AEs during 28 days after each vaccination; and occurrence of medically attended AEs, serious AEs, and AEs of special interest throughout the 7-month study period. Induction of neutralising antibody measured by PNA was expressed as a geometric mean titer (GMT) at 14days post second vaccination, a GMT ratio in subjects seronegative at baseline, a geometric mean fold rise (GMFR), and a percentage of subjects with a ≥4-fold increase from baseline regardless of baseline anti-spike IgG seropositivity. A secondary immunogenicity outcome was the induction of anti-spike IgG in binding antibody units (BAU/mL) expressed in the same four parameters used for the neutralising activity. The exploratory immunogenicity outcomes were the induction of neutralising antibodies to wild-type SARS-CoV-2 expressed as a GMT and GMT ratio (COVIVAC/VAXZEVRIA) at 14days post second vaccination, and the frequency of spike-specific activated T cells.

### Statistical analysis

This study (ClinicalTrials.gov NCT05940194) was descriptive with no confirmatory objective, as it was intended to assess the feasibility of advancing the evaluation of COVIVAC towards emergency use authorization based on superiority to the comparator. The study had >90% power to demonstrate a lower bound of the 95% confidence interval (CI) of the GMT ratio greater than 1·0 if the observed ratio (COVIVAC/VAXZEVRIA) was ≥1·65. The study also had >95% power to detect at least one serious or severe adverse event if the underlying rate was ≥2·5% and power was >80% to detect differences in AE rates ≥15%. All statistical tests were two-sided with a significance level of 0·05. All statistical analyses were performed using SAS version 9·4. All safety assessments occurred in the treatment-exposed population, according to the treatment received. All treatment group percentages were supplemented with two-sided 95% confidence intervals (CIs) computed via the Clopper-Pearson method. The immunogenicity analysis presented was performed in the full analysis population that included all subjects randomised for whom any post-vaccination immunogenicity data were available. This population is identical to the per-protocol population at Day 43. Geometric mean antibody responses were reported by treatment and time point, accompanied by 95% CIs. The analysis of geometric means excluded subjects who were seropositive at baseline (defined by anti-spike IgG >LLOQ as measured by ELISA). Geometric mean fold rises (GMFR) were calculated relative to baseline using the log difference of the paired samples, with corresponding CIs computed via the t-distribution, utilizing the antilog transformation to present the ratio. The proportions of subjects with GMFRs of NT50 ≥4 from baseline were summarized with two-sided 95% confidence intervals computed via the Clopper-Pearson method. The analysis of immunogenicity relative to baseline included baseline seropositive subjects. The CD4+ T cell responses were summarized via the geometric mean and treatment groups were compared via the Mann-Whitney U test.

## Results

From August 10 to 23, 2021, 737 individuals were screened and 374 were randomised to three treatment groups (124-125 subjects per group). All received dose one; three missed dose two; 365 completed the last study visit on day 197 (Figure 1). The baseline characteristics are shown by treatment group in Table 1; the exposed population was 49·5% male, had a mean age of 49 years (range 18-77) and a mean body mass index of 22·29 (range 17·01-31·76).

**Figure 1.**
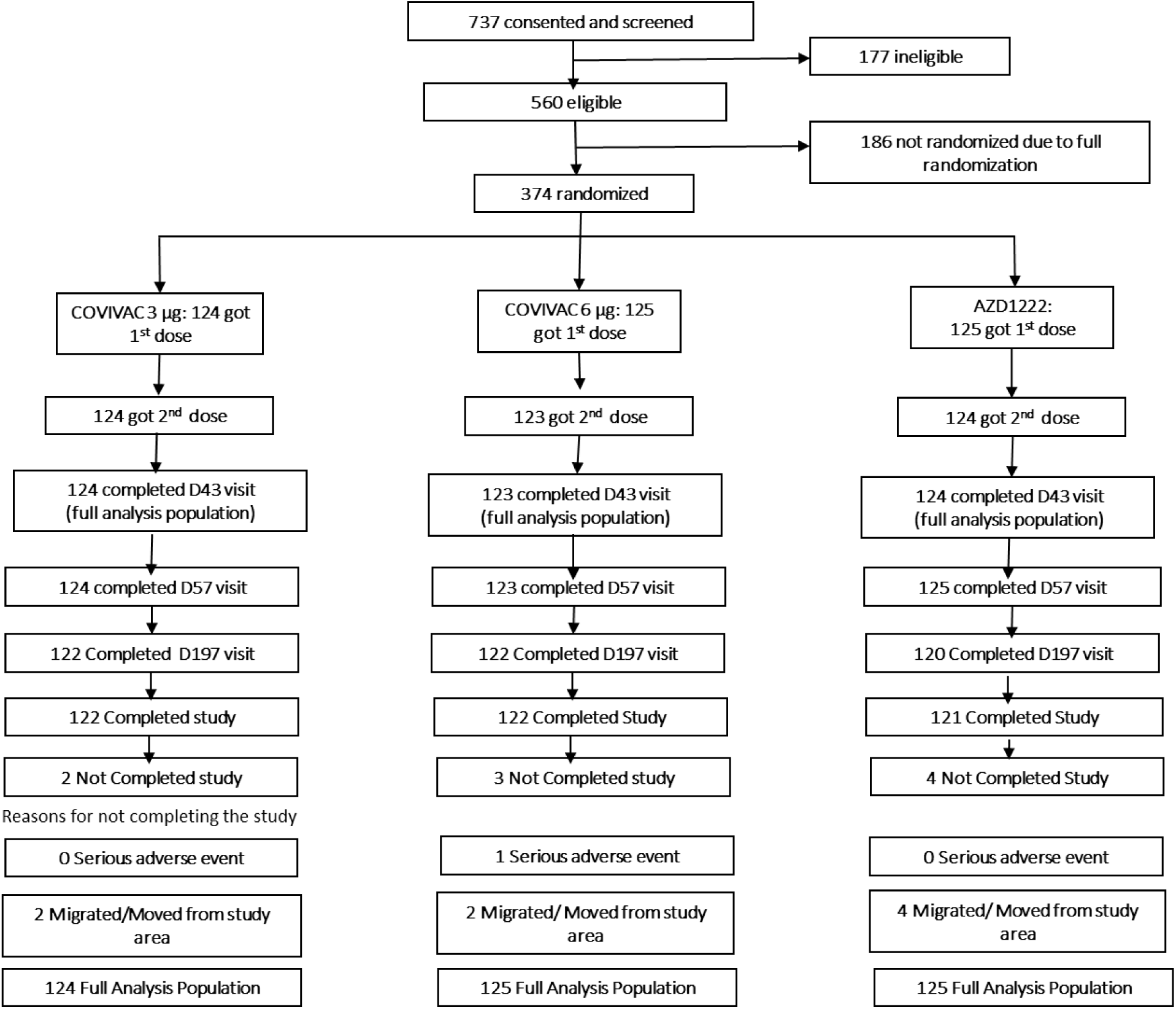
Cohort disposition: Disposition of subjects recruited and randomized in Phase 2

**Table 1:** Baseline characteristics of the exposed population.

| Baseline Characteristics | COVIVAC |  | VAXZEVRIA |
| --- | --- | --- | --- |
|  | 3 µg (N = 124) | 6 µg (N = 125) | (N = 125) |
| Mean age in years (SD; range) | 48·9 (14·82; 18, 75) | 48·9 (14·27; 18,77) | 49·8 (14·17; 18,74) |
| Male | 64 (51·6%) | 67 (53·6%) | 54 (43·2%) |
| Female | 60 (48·4%) | 58 (46·4%) | 71 (56·8%) |
| Mean body mass index in kg/m2 (SD; range) | 22·29<br>(2·57; 17·01, 31·76) | 22·27<br>(2·67; 17·33, 28·75) | 22·24<br>(2·55; 17·08, 28·88) |

Trial participants from all three vaccine groups tolerated the doses with no dose-limiting reactogenicity. Solicited injection site reactogenicity was mostly mild during the seven days after each vaccination (Table 2). Pain or tenderness was the most common injection site symptom recorded, more frequently following dose one than dose two. Post-dose one-injection site pain was reported by 72% of VAXZEVRIA recipients but by only 46–56% of COVIVAC recipients. The most common systemic symptoms (Table 2) were fatigue or malaise, headache, and myalgia, reported more frequently following dose one than dose two. Notably, fever (≥38°C) following dose one occurred in 22·4% of VAXZEVRIA recipients but in only 0·8% of COVIVAC recipients. Unsolicited adverse events occurring 28 days after vaccination (Supplement Table 2) were reported by a similar proportion of subjects in each treatment group (23·2–31·2%); none of these events were judged by the investigator to be treatment-related or led to withdrawal from the trial. Although six serious adverse events were reported during the entire study period (three in each COVIVAC treatment group), none were considered treatment-related (intestinal obstruction, sialadenitis, leukemia, COVID-19, colon cancer, and gastric cancer). The independent DSMB expressed no safety concerns.

**Table 2:** Number of subjects with solicited adverse events during 7 days after vaccination in the safety analysis population.

| Reaction |  | COVIVAC |  | VAXZEVRIA |
| --- | --- | --- | --- | --- |
|  |  | 3 µg (N = 124)<br>n (%) (95% CI*) | 6 µg (N = 125)<br>n (%) (95% CI*) | (N = 125)<br>n (%) (95% CI*) |
| Pain/tenderness | Dose 1 | 57 (46·0%) | 65 (52·0%) | 90 (72·0%) |
|  |  | (37·0-55·1) | (42·9-61·0) | (63·3-79·7) |
|  | Dose 2 | 34 (27·4%) | 48 (39·0%) | 37 (29·8%) |
|  |  | (19·8-36·2) | (30·4-48·2) | (22·0-38·7) |
| Swelling/induration | Dose 1 | 2 (1·6%) | 1 (0·8%) | 1 (0·8%) |
|  |  | (0·2-5·7) | (0·0-4·4) | (0·0-4·4) |
|  | Dose 2 | 2 (1·6%) | 0 (0%) | 0 (0%) |
|  |  | (0·2-5·7) | (0·0-3·0) | (0·0-2·9) |
| Erythema | Dose 1 | 0 (0%) | 0 (0%) | 1 (0·8%) |
|  |  | (0·0-2·9) | (0·0-2·9) | (0·0-4·4) |
|  | Dose 2 | 1 (0·8%) | 0 (0%) | 0 (0%) |
|  |  | (0·0-4·4) | (0·0-3·0) | (0·0-2·9) |
| Fever (≥ 38 °C) | Dose 1 | 1 (0·8%) | 1 (0·8%) | 28 (22·4%) |
|  |  | (0·0-4·4) | (0·0-4·4) | (15·4-30·7) |
|  | Dose 2 | 3 (2·4%) | 3 (2·4%) | 2 (1·6%) |
|  |  | (0·5-6·9) | (0·5-7·0) | (0·2-5·7) |
| Headache | Dose 1 | 31 (25·0%) | 52 (41·6%) | 63 (50·4%) |
|  |  | (17·7-33·6) | (32·9-50·8) | (41·3-59·5) |
|  | Dose 2 | 28 (22·6%) | 28 (22·8%) | 26 (21·0%) |
|  |  | (15·6-31·0) | (15·7-31·2) | (14·2-29·2) |
| Fatigue/malaise | Dose 1 | 53 (42·7%) | 59 (47·2%) | 77 (61·6%) |
|  |  | (33·9-51·9) | (38·2-56·3) | (52·5-70·2) |
|  | Dose 2 | 39 (31·5%) | 38 (30·9%) | 40 (32·3%) |
|  |  | (23·4-40·4) | (22·9-39·9) | (24·1-41·2) |
| Myalgia | Dose 1 | 24 (19·4%) | 26 (20·8%) | 47 (37·6%) |
|  |  | (12·8-27·4) | (14·1-29·0) | (29·1-46·7) |
|  | Dose 2 | 21 (16·9%) | 14 (11·4%) | 23 (18·5%) |
|  |  | (10·8-24·7) | (6·4-18·4) | (12·1-26·5) |
| Arthralgia | Dose 1 | 23 (18·5%) | 17 (13·6%) | 31 (24·8%) |
|  |  | (12·1-26·5) | (8·1-20·9) | (17·5-33·3) |
|  | Dose 2 | 5 (4·0%) | 9 (7·3%) | 16 (12·9%) |
|  |  | (1·3-9·2) | (3·4-13·4) | (7·6-20·1) |
| Nausea/vomiting | Dose 1 | 10 (8·1%) | 9 (7·2%) | 13 (10·4%) |
|  |  | (3·9-14·3) | (3·3-13·2) | (5·7-17·1) |
|  | Dose 2 | 2 (1·6%) | 4 (3·3%) | 3 (2·4%) |
|  |  | (0·2-5·7) | (0·9-8·1) | (0·5-6·9) |

The main immunogenicity measure was the induction of vaccine-homologous antibodies assessed by PNA 14 days after vaccine dose two. Figure 2 shows plots of neutralising (PNA) antibody GMT by age and treatment group over time among the 95% of subjects seronegative at baseline for anti-S IgG and with a valid assay result (see also Supplement Table 3). Responses to COVIVAC were significantly higher than to VAXZEVRIA 14 days after vaccine dose two, although this contrast was not statistically significant six months after vaccine dose two. Note that six months after dose two, GMTs remained well above baseline, with increases in two groups among adults 18-59 years of age, presumably due to intercurrent infection with SARS-CoV-2. The percentage of subjects 18-59 years of age mounting a minimum four-fold PNA response to vaccination 14 days after vaccine dose two was 89·0% (95% CI 80·2-94·9) for COVIVAC 3µg, 92·8% (95% CI 84·9-97·3) for COVIVAC 6µg, and 85·4% (95% CI 75·8-92·2) for VAXZEVRIA. Equally high PNA response rates were also observed in COVIVAC vaccinees ≥60 years of age (Supplement Table 4). Notably, the magnitude of neutralising antibody induction 14 days after dose two, expressed as a PNA GMFR from baseline, although similar between COVIVAC groups, was greater compared to the VAXZEVRIA group (Table 3) for subjects 18-59 years of age and for subjects ≥60 years of age. The greater peak induction of neutralising antibodies by COVIVAC relative to VAXZEVRIA was also apparent in the GMT ratios (COVIVAC/VAXZEVRIA) for both dose levels with 95% confidence intervals that excluded 1·00 for both age strata (Supplement Table 5).

**Figure 2.**
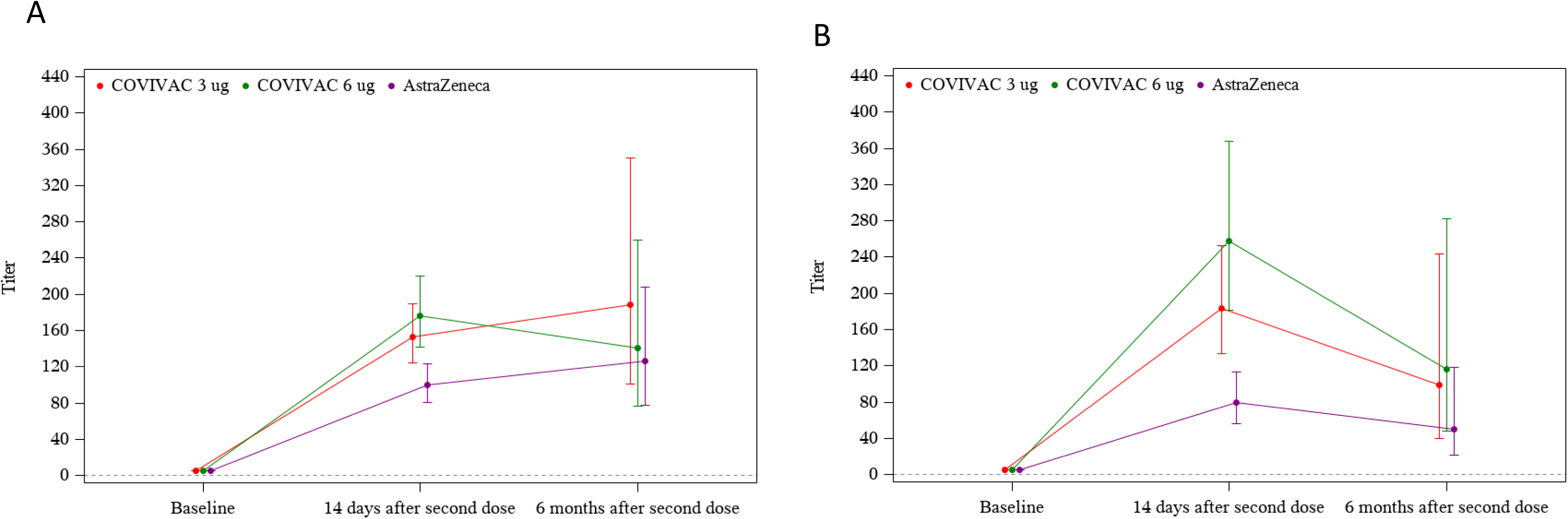
Geometric mean titer and 95% CI of NT50 against SARS-CoV-2 pseudotyped virus by age and treatment group in the full analysis population: (a) 18-59 years, (b) ≥ 60 years

**Table 3:**
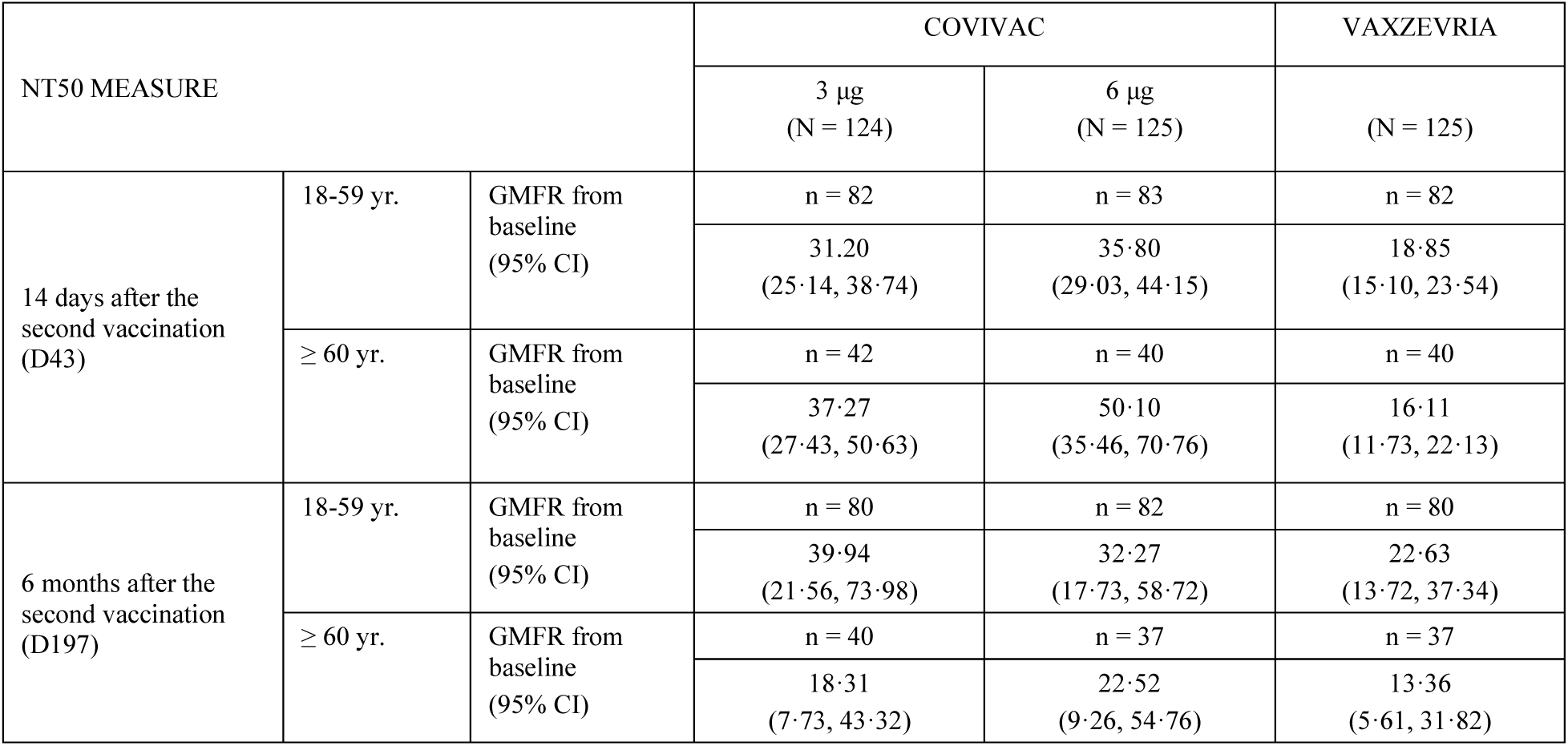
Summary of geometric mean fold rise (GMFR) from baseline of NT50 titers against SARS-CoV-2 pseudovirus by age group in the full analysis population.

| NT50 MEASURE |  |  | COVIVAC |  | VAXZEVRIA |
| --- | --- | --- | --- | --- | --- |
|  |  |  | 3 µg<br>(N = 124) | 6 µg<br>(N = 125) | (N = 125) |
| 14 days after the second vaccination (D43) | 18-59 yr. | GMFR from baseline (95% CI) | n = 82 | n = 83 | n = 82 |
|  |  |  | 31.20<br>(25.14, 38.74) | 35.80<br>(29.03, 44.15) | 18.85<br>(15.10, 23.54) |
|  | ≥ 60 yr. | GMFR from baseline (95% CI) | n = 42 | n = 40 | n = 40 |
|  |  |  | 37.27<br>(27.43, 50.63) | 50.10<br>(35.46, 70.76) | 16.11<br>(11.73, 22.13) |
| 6 months after the second vaccination (D197) | 18-59 yr. | GMFR from baseline (95% CI) | n = 80 | n = 82 | n = 80 |
|  |  |  | 39.94<br>(21.56, 73.98) | 32.27<br>(17.73, 58.72) | 22.63<br>(13.72, 37.34) |
|  | ≥ 60 yr. | GMFR from baseline (95% CI) | n = 40 | n = 37 | n = 37 |
|  |  |  | 18.31<br>(7.73, 43.32) | 22.52<br>(9.26, 54.76) | 13.36<br>(5.61, 31.82) |

To confirm the observation of COVIVAC’s superior peak induction of vaccine-homologous neutralizing antibodies, we evaluated the GMT of neutralising antibodies measured by live virus neutralizing assay (LVNA) induced by two doses of 3µg or 6µg of COVIVAC and compared it with neutralising antibodies induced by VAXZEVRIA. Although the GMTs measured by live virus assays were approximately two thirds less than mean titers measured by pseudotyped virus, the GMT ratios estimated for the two COVIVAC groups relative to the VAXZEVRIA group by either assay were 1.5 to 2.0-fold higher in a dose-dependent manner (Supplement Table 6).

A secondary immunogenicity outcome was the induction of anti-spike IgG in binding antibody units (BAU/mL). By this measure of immunogenicity, VAXZEVRIA induced higher peak concentrations of anti-spike IgG measured by ELISA than did COVIVAC at either dose or for both age strata (Supplement Tables 7–9). For instance, the GMC ratio (COVIVAC 3µg/VAXZEVRIA) at 14 days after vaccine dose two in subjects 18-59 was 0·38 (95% CI, 0·29-0·50), and in those ≥60 was 0·47 (95% CI 0·28-0·78). Six months after dose two, the 95% CI for the GMC ratios included 1·00. This aligns with earlier observations showing that inactivated NDV-HXP S induces higher neutralising antibody-to-spike binding antibody ratios compared to other vaccine platforms.^19^

Finally, we explored the induction of spike specific CD4+ T cell responses by COVIVAC and VAXZEVRIA in a random subset of vaccinated individuals with no detectable anti-spike IgG by ELISA at baseline. Spike specific CD4+ T cell response was assessed utilizing an activation-induced molecules (AIM) assay, which evaluates the frequency of antigen-specific T cells based on the co-expression of OX40 and CD137 for CD4+ T cells and CD69 and CD137 of CD8+T cells (Figure 3). We detected induction of a spike specific CD4+ T cell response on day 43 in all of 10 COVIVAC 3 μg vaccinees with a 0·14% cell frequency (95% CI 0·074-0·27%), in 9 of 10 COVIVAC 6 μg vaccinees with a 0·092% cell frequency (95% CI 0·040-0·21%), and in all of 12 VAXZEVRIA vaccinees with a 0·18% cell frequency (95% CI 0·095-0·34%) (Figure 3B). The intensity of spike specific CD4+ T cell induction on day 43 was similar among the treatment groups (Figure 3C).

**Figure 3.**
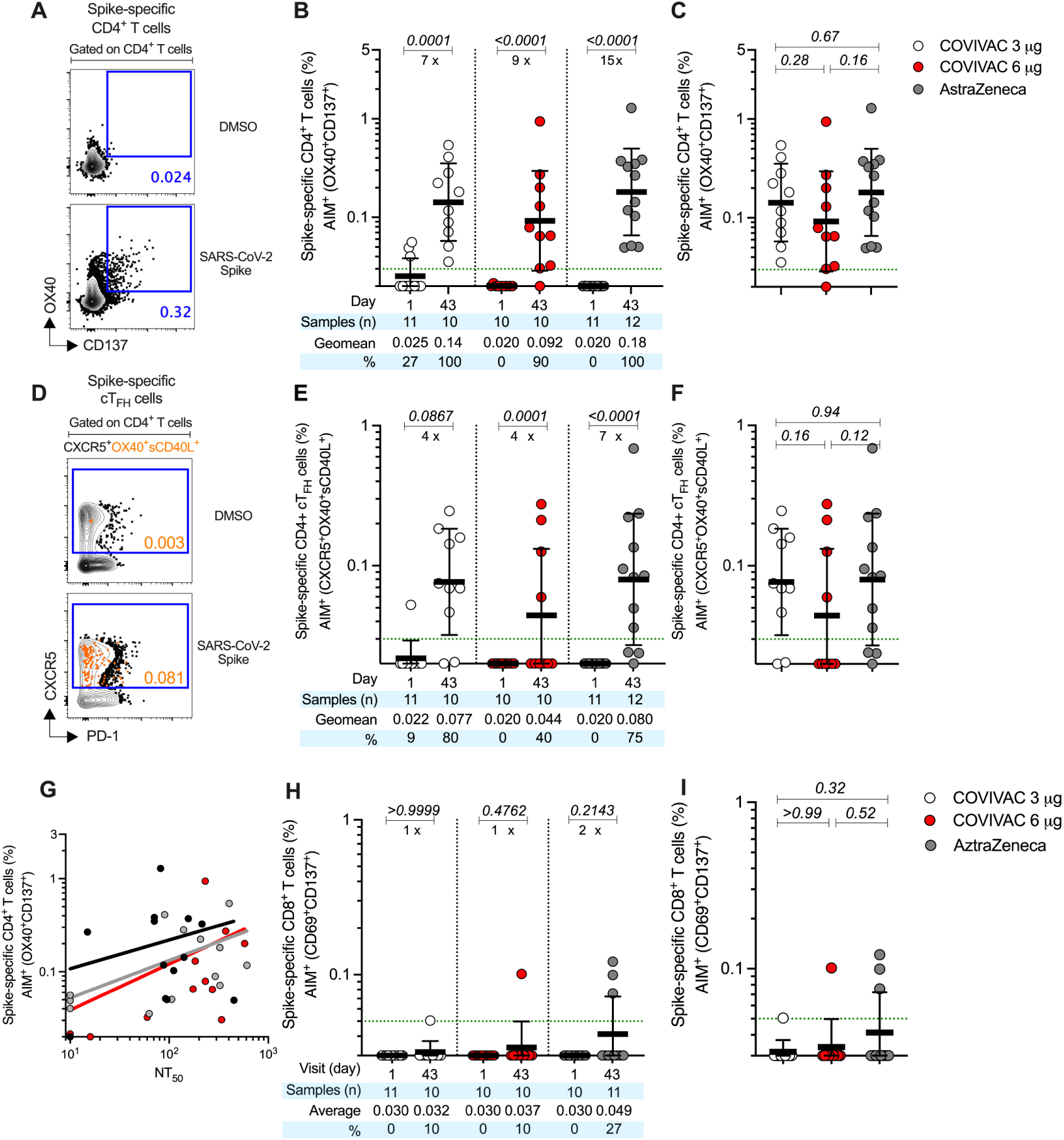
SARS-CoV-2 spike-specific CD4+ T cell responses are induced by COVIVAC. (A) FACS example of SARS-CoV-2 spike-specific CD4+ T cells evaluated by the AIM assay after stimulation with spike MP. Spike-specific CD4+ T cells were quantified by AIM (surface OX40+CD137+) after stimulation with spike peptide megapool (MP). (B) Frequencies of SARS-CoV-2 spike-specific CD4+ T cells induced by COVIVAC at 3 µg and 6 µg and VAXZEVRIA at day 1 (baseline) and at day 43 post-vaccination. (C) Comparison of spike specific CD4+ T cells induced by COVIVAC at 3 µg and 6 µg and VAXZEVRIA at 43 days post-vaccination. (D) FACS example of SARS-CoV-2 spike-specific circulating follicular helper T (cTFH) cells (CXCR5+OX40+surface CD40L+, as a percentage of CD4+ T cells) after stimulation with spike MP. (E) Frequencies of SARS-CoV-2 spike-specific cTFH cells induced by COVIVAC at 3 µg and 6 µg and VAXZEVRIA at day 1 (baseline) and at day 43 post-vaccination. (F) Comparison of spike-specific cTFH cells induced by COVIVAC at 3 µg and 6 µg and VAXZEVRIA at 43 days post-vaccination. Dotted green lines indicate the limit of quantification (LOQ). Light gray, COVIVAC at 3 µg; red, COVIVAC at 6 µg; black, VAXZEVRIA. G) Correlation of spike specific CD4 T cell responses and neutralizing antibody titers measured 43 days post-vaccination for COVIVAC at 3 µg (grey line) and 6 µg (red line) and VAXZEVRIA (black line). H) Comparison of spike specific CD8+ T cells induced by COVIVAC at 3 µg and 6 µg and VAXZEVRIA at 43 days post-vaccination. I) Comparison of spike specific CD8+ T cells s induced by COVIVAC at 3 µg and 6 µg and VAXZEVRIA at 43 days post-vaccination. The bars in (B, C, E, F, H, I) indicate the geometric mean and geometric SD in the analysis of the spike-specific T cell frequencies. Data were analysed for statistical significance using the Mann-Whitney U test (B, C, E, F, H, I). Background-subtracted and log data analysed in all cases.

Follicular helper T (TFH) cells help B cells activate antibody production. As this T cell subset can be induced by SARS-CoV-2 infection and COVID-19 vaccination, we evaluated the frequency of circulating TFH cells by the AIM assay at baseline (day 1) and post-dose two (day 43) (Figure 3E). cTFH were detected on day 43 in 8 of 10 COVIVAC 3 μg vaccinees with a 0·077% cell frequency (95% CI 0·020-0·097%), in 4 of 10 COVIVAC 6 μg vaccinees with a 0·044% cell frequency (95% CI 0·041-0·14%), and in 9 of 12 VAXZEVRIA vaccinees with a 0.08% cell frequency (95% CI 0·040-0·16%) (Figure 3E). As shown for the spike specific CD4+ T cells, the intensity of spike specific cTFH cells induction on day 43 was similar among treatment groups (Figure 3F).

Antibody levels by PNA and frequencies of memory CD4+ T cells were significantly correlated for COVIVAC 3 ug (r=0.824, p>0.0001), COVIVAC 6 ug (r=0.875, p>0.0001), and VAXZEVRIA (r=0.764, p>0.0001) (Figure 3G); this finding is evidence of a coordinated cellular-humoral immune response in both COVIVAC and VAXZEVRIA recipients.

Spike specific CD8+ T cells were also measured by AIM (CD69+ CD137+). We detected a response on day 43 in 1 of 10 COVIVAC 3 μg vaccinees with a 0·032% cell frequency (95% CI 0·0.028-0·0.036%), in 1 of 10 COVIVAC 6 μg vaccinees with a 0·037% cell frequency (95% CI 0.023-0.052%), and in 3 of 11 VAXZEVRIA vaccinees with a 0·049% cell frequency (95% CI 0.029-0.069%) (Figure 3H). The intensity of CD8+ T cell responses detected on day 43 was low and similar among the treatment groups. (Figure 3I).

## Discussion

This phase 2 study showed that COVIVAC (NDV-HXP-S), when administered as a two-dose series to adults, including those 60 years of age and older, has an acceptable safety profile. It is highly immunogenic, activating T cell responses, and eliciting neutralising antibody responses 14 days after vaccine dose two that are superior to those induced by the adenovirus vectored VAXZEVRIA comparator vaccine.

All treatments evaluated were well tolerated with predominantly mild and self-limited reactogenicity that was greater after dose one than after dose two. The COVIVAC formulations at 3 and 6 µg dose levels were less reactogenic after dose one than the VAXZEVRIA comparator with respect to self-reported pain at the injection site, myalgia, and incidence of fever. Otherwise, there were no notable differences. Overall, in this study of 374 participants, there were no spontaneously reported AEs attributed by investigators to vaccination and no concerns expressed by the DSMB providing safety oversight.

In terms of neutralising antibody titers, measured in a PNA, both dose levels of COVIVAC showed superiority to VAXZEVRIA within each age stratum at an early time point (14 days after dose two) with the trend continuing out to month 6, even though statistical significance was not reached at the later time point. Superior induction of neutralizing antibody by COVIVAC at both dose levels relative to VAXZEVRIA 14 days after vaccine dose two was confirmed by exploratory testing using a live virus neutralization assay. Interestingly, spike-binding antibodies were lower in the COVIVAC groups compared to the VAXZEVRIA group, hinting at a better ratio of neutralising to binding antibodies for COVIVAC. In fact, it has been shown in an earlier study, that inactivated NDV-HXP-S vaccines induce better ratios of neutralising antibodies relative to spike binding IgG compared to mRNA vaccines.^19^ These findings are important since neutralising antibodies to SARS-CoV-2 are a mechanistic correlate of protection^5,20^ and new SARS-CoV-2 vaccines (e.g. Corbevax and VLA2001) have been licensed based on immune-bridging of neutralising antibody titers.^21-22^ Our results suggest that the possibility could be open for COVIVAC or for similar NDV-based vaccines to be developed by other manufacturers.

There was no improvement in the induction of vaccine homologous neutralising antibodies by doubling the dose from a 3 to 6 µg level. Considering the important dose effect on immunogenicity observed in the phase 1 trial comparing 10 and 3 µg dose levels, with no adverse impact on reactogenicity, further development of COVIVAC will likely revert to a 10 µg dose level.^8^

In this comparative study, spike specific CD4+ T cell responses were detected in 90-100% of a small subset of randomly selected individuals, all being seronegative for anti-spike IgG pre-vaccination, in test and comparator vaccine groups. This is comparable to what has been reported for other COVID-19 vaccines such as mRNA and adenovirus vector vaccines.^14^ Similarly, we have detected circulating T follicular helper cells in a substantial fraction of vaccinees, supported by the strong correlation of spike-specific CD4+ T cell responses and functional antibody responses. In previous studies, we demonstrated that a coordinated cellular-humoral immune response is associated with mild disease outcomes in infected individuals.^14,18^.

This study has several limitations. First, it was a phase 2 trial of limited size with no clinical endpoint. Second, the investigational and comparator vaccines expressed an ancestral spike immunogen. Moreover, the study population was largely naïve to SARS-CoV-2 at the time they were vaccinated. Currently, COVID-19 vaccines are being deployed for booster immunization in primed butat risk adults. While COVIVAC performed well by inducing neutralising antibodies, its use as a booster vaccine is yet to be evaluated. Vaccines with ancestral spike antigens are obsolete now due to emergence of different variants, especially the Omicron variant family. Current recommendations from regulatory authorities and WHO state that XBB-lineage spike antigens should be used in updated vaccines. GMP seed viruses for COVIVAC with XBB.1.5 spike exist and can be used for manufacturing of strain-changed updated vaccines. We did not evaluate induction of neutralising antibodies to vaccine heterologous variants, as this was outside of the scope of this study. Nevertheless, we observed that COVIVAC induced CD4+ T cell responses comparable to the VAXZEVRIA comparator, and it has been reported that CD4+ T cell responses induced by the ancestral spike protein are maintained and cross-recognize SARS-CoV-2 variants, from Alpha to Omicron.^23,24^

Strengths of this study are the use of a fully validated functional antibody readout (PNA), the inclusion of older adults with an age-stratified analysis showing preservation of immunogenicity despite increased age, the assessment of T cell responses, and the selection of the VAXZEVRIA vaccine as a highly relevant immuno-bridging comparator. The efficacy of the VAXZEVRIA vaccine has been demonstrated in multiple double-blind randomized clinical trials, varying from approximately 70% against any symptomatic disease to >95% against severe disease and/or hospitalization.^25^ Multiple effectiveness and observational studies confirmed the high level of protection afforded by the vaccine, leading to its approval in the UK and other European countries.^26^ By early 2022, the VAXZEVRIA vaccine had been approved by over 170 countries, including Vietnam, making it the most widely deployed vaccine across the globe with over 2.5 billion doses used.^26^ The induction of superior levels of neutralizing antibodies by COVIVAC and similar activation of CD4+ T cells in comparison to VAXZEVRIA strongly suggest that COVIVAC’s effectiveness would be at least similar.

The CD4+ and CD8+ T cell response has been assessed using the AIM assay measuring the frequency of spike-specific T cell responses. It is important to point out that functional capacity of T cell responses, such as through production of cytokines, need to be assessed for a comprehensive picture of vaccine-induced spike-specific T-cell responses. ^27^

The clinical trial was designed to assess the feasibility of conducting a phase 3 trial in which the benefit of vaccination with COVIVAC could be confirmed by demonstrating non-inferior or superior immunogenicity relative to an authorized comparator COVID-19 vaccine. That aim was met. Further development of COVIVAC updated to express a contemporary recombinant spike protein, administered as a booster dose to vulnerable individuals, is a viable option for its manufacturer IVAC, which serves the public sector of Vietnam.

## Data Availability

All data produced in the present work are contained in the manuscript

## Contributors

VDT and LDM verified the underlying data reported herein. All authors had full access to all the data in the study and had final responsibility for the decision to submit for publication. Individual author roles are reported using CRediT: Conceptualisation, TCT, NMH, LDM, DHT, LVB, BLI, FK, DW; Data curation, VDT, DDA, VHH, NVT, TCT, JM, JMC, NMH, LDM, JF, DHT, DW; Formal analysis, LDM; Funding acquisition, RR, LVB, AS, BLI, FK, DW; Investigation, VDT, DDA, VHH, NVT, JM, JMC, EAE, AR, DW; Methodology, RR, LDM, JF, FK, DW; Project administration, TCT, NMH, DHT, LVH; Resources, VDT, DDA, NVT, AS, FK, DW; Supervision, VDT, DDA, JM, JMC, RR, BLI, FK, DW; Validation, LDM; Visualisation, BLI; Writing-original draft, RR, BLI, FK, DW; Writing−review & editing, VDT, DDA, VHH, NVT, TCT, JM, JMC, NMH, LDM, JF, EAE, AR, DHT, LVB, AS.

## Data Sharing

The study protocol is provided in the supplement. Deidentified participant data will be made available for two years after publication upon request directed to the lead author Vu Dinh Thiem. After approval of a data-sharing proposal, data can be shared through a secure online platform.

## Acknowledgments

Work at Mount Sinai was supported by philanthropic donations to Mount Sinai/institutional funding (C-VaRPP funding). Preclinical development of the COVIVAC vaccine was supported, in part, by the Bill & Melinda Gates Foundation [INV-021239]. Under the grant conditions of the Foundation, a Creative Commons Attribution 4.0 Generic License has already been assigned to the Author Accepted Manuscript version that might arise from this submission. Initial work on COVIVAC was also supported by institutional funding from the Icahn School of Medicine at Mount Sinai. This work has been partially supported by the National Institutes of Health under Contract No. 75N93019C00065 to A.S, D.W. PATH was also supported by the Coalition for Epidemic Preparedness and a charitable contribution from Bayer AG. We thank the University of Texas at Austin for granting IVAC access to their 6-proline stabilized SARS-CoV-S (Hexapro) technology.

## Figure legends

**Figure S1:**
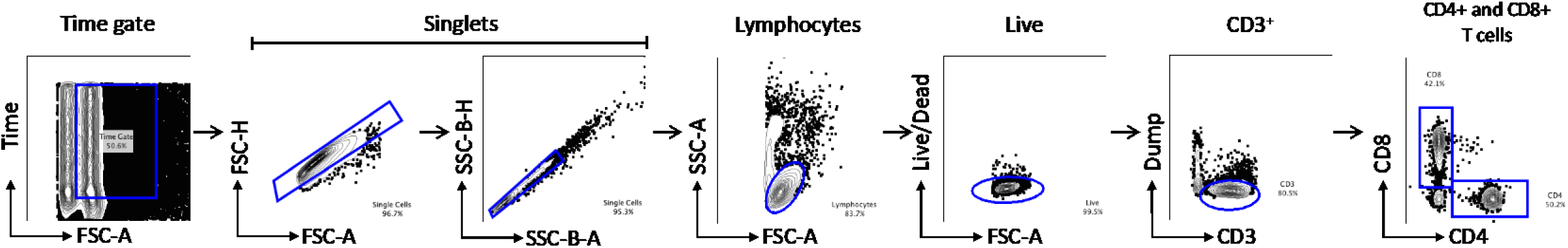
Gating strategy to identify CD4 and CD8 T lymphocytes: Representative gating strategy to define CD3+CD4+ and CD3+CD8+ cells by AIM assay.

## References

1. Duroseau B, Kipshidze N, Limaye RJ. The impact of delayed access to COVID-19 vaccines in low- and lower-middle-income countries. Front Public Health. 2022; 10:1087138.

2. Ministry of Health portal. COVID-19 epidemic prevention bulletin dated 3/1 of the Ministry of Health. Ministry of Health [Internet]. 2023 Mar 1 [cited 2023 Jun 27]; Available from: https://moh.gov.vn/tin-tong-hop/-/asset_publisher/k206Q9qkZOqn/content/ban-tin-phong-chong-dich-covid-19-ngay-3-1-cua-bo-y-te

3. World Health Organization. WHO releases global COVID-19 vaccination strategy update to reach unprotected. World Health Organization [Internet]. 2022 Jul 22; Available from: https://www.who.int/news/item/22-07-2022-who-releases-global-covid-19-vaccination-strategy-update-to-reach-unprotected

4. Khoury DS, Cromer D, Reynaldi A, Schlub TE, Wheatley AK, Juno JA, et al. Neutralizing antibody levels are highly predictive of immune protection from symptomatic SARS-CoV-2 infection. Nat Med. 2021 Jul;27(7):1205–11.

5. Gilbert PB, Montefiori DC, McDermott AB, Fong Y, Benkeser D, Deng W, et al. Immune correlates analysis of the mRNA-1273 COVID-19 vaccine efficacy clinical trial. Science. 2022 Jan 7;375(6576):43–50.

6. Tarke A, Coelho CH, Zhang Z, Dan JM, Yu ED, Methot N, et al. SARS-CoV-2 vaccination induces immunological T cell memory able to cross-recognize variants from Alpha to Omicron. Cell. 2022 Mar 3;185(5):847–859.e11.

7. Sun W, Liu Y, Amanat F, González-Domínguez I, McCroskery S, Slamanig S, et al. A Newcastle disease virus expressing a stabilized spike protein of SARS-CoV-2 induces protective immune responses. Nat Commun. 2021 Oct 27;12(1):6197.

8. Duc Dang A, Dinh Vu T, Hai Vu H, Thanh Ta V, Thi Van Pham A, Thi Ngoc Dang M, et al. Safety and immunogenicity of an egg-based inactivated Newcastle disease virus vaccine expressing SARS-CoV-2 spike: Interim results of a randomized, placebo-controlled, phase 1/2 trial in Vietnam. Vaccine. 2022 Jun 9;40(26):3621–32.

9. Voysey M, Clemens SAC, Madhi SA, Weckx LY, Folegatti PM, Aley PK, et al. Safety and efficacy of the ChAdOx1 nCoV-19 vaccine (AZD1222) against SARS-CoV-2: an interim analysis of four randomised controlled trials in Brazil, South Africa, and the UK. The Lancet. 2021 Jan;397(10269):99–111.

10. Bewley KR, Coombes NS, Gagnon L, McInroy L, Baker N, Shaik I, et al. Quantification of SARS-CoV-2 neutralizing antibody by wild-type plaque reduction neutralization, microneutralization and pseudotyped virus neutralization assays. Nat Protoc. 2021 Jun;16(6):3114–40.

11. Carreño JM, Alshammary H, Singh G, Raskin A, Amanat F, Amoako A, et al. Evidence for retained spike-binding and neutralizing activity against emerging SARS-CoV-2 variants in serum of COVID-19 mRNA vaccine recipients. EBioMedicine. 2021 Nov;73:103626.

12. Havenar-Daughton C, Reiss SM, Carnathan DG, Wu JE, Kendric K, Torrents de la Peña A, et al. Cytokine-Independent Detection of Antigen-Specific Germinal Center T Follicular Helper Cells in Immunized Nonhuman Primates Using a Live Cell Activation-Induced Marker Technique. The Journal of Immunology. 2016 Aug 1;197(3):994–1002.

13. Reiss S, Baxter AE, Cirelli KM, Dan JM, Morou A, Daigneault A, et al. Comparative analysis of activation induced marker (AIM) assays for sensitive identification of antigen-specific CD4 T cells. PLoS One. 2017 Oct 24;12(10):e0186998.

14. Zhang Z, Mateus J, Coelho CH, Dan JM, Moderbacher CR, Gálvez RI, et al. Humoral and cellular immune memory to four COVID-19 vaccines. Cell. 2022 Jul 7;185(14):2434–2451.e17.

15. Ogbunude POJ. Efflux of 3H-thymidine by erythrocytes from mice infected with Trypanosoma brucei brucei. Ann Trop Med Parasitol. 1986 Dec 15;80(6):581–5.

16. Dan JM, Mateus J, Kato Y, Hastie KM, Yu ED, Faliti CE, et al. Immunological memory to SARS-CoV-2 assessed for up to 8 months after infection. Science. 2021 Feb 5;371(6529).

17. Mateus J, Dan JM, Zhang Z, Rydyznski Moderbacher C, Lammers M, Goodwin B, et al. Low-dose mRNA-1273 COVID-19 vaccine generates durable memory enhanced by cross-reactive T cells. Science. 2021 Oct 22;374(6566):eabj9853.

18. Grifoni A, Weiskopf D, Ramirez SI, Mateus J, Dan JM, Moderbacher CR, et al. Targets of T Cell Responses to SARS-CoV-2 Coronavirus in Humans with COVID-19 Disease and Unexposed Individuals. Cell. 2020 Jun;181(7):1489–1501.e15.

19. Carreño JM, Raskin A, Singh G, Tcheou J, Kawabata H, Gleason C, et al. An inactivated NDV-HXP-S COVID-19 vaccine elicits a higher proportion of neutralizing antibodies in humans than mRNA vaccination. Sci Transl Med. 2023 Feb 15;15(683).

20. Goldblatt D, Fiore-Gartland A, Johnson M, Hunt A, Bengt C, Zavadska D, et al. Towards a population-based threshold of protection for COVID-19 vaccines. Vaccine. 2022 Jan;40(2):306–15.

21. European Medicines Agency. COVID-19 Vaccine (inactivated, adjuvanted) Valneva [Internet]. 2022. Available from: https://www.ema.europa.eu/en/medicines/human/EPAR/covid-19-vaccine-inactivated-adjuvanted-valneva#safety-updates-section

22. Biological E. Limited. Summary of Product Characteristics (SmPC) SARS-CoV-2 (Covid-19) Vaccine CORBEVAX^TM^ [Internet]. 2022 Feb. Available from: https://cdsco.gov.in/opencms/resources/UploadCDSCOWeb/2018/UploadSmPC/ebio.pdf

23. Grifoni A, Sette A. From Alpha to omicron: The response of T cells. Current Research in Immunology. 2022;3:146–50.

24. Petrone L, Sette A, de Vries RD, Goletti D. The Importance of Measuring SARS-CoV-2-Specific T-Cell Responses in an Ongoing Pandemic. Pathogens. 2023 Jun 22;12(7):862.

25. Voysey M, Costa Clemens SA, Madhi SA, Weckx LY, Folegatti PM, Aley PK, et al. Single-dose administration and the influence of the timing of the booster dose on immunogenicity and efficacy of ChAdOx1 nCoV-19 (AZD1222) vaccine: a pooled analysis of four randomised trials. The Lancet. 2021 Mar;397(10277):881–91.

26. United Kingdom Department of Health and Social Care, Javid S, Troup M. One year anniversary of UK deploying Oxford-AstraZeneca vaccine. 2022 Jan 4; Available from: https://www.gov.uk/government/news/one-year-anniversary-of-uk-deploying-oxford-astrazeneca-vaccine

27. Zhang Z, Mateus J, Coelho CH, Dan JM, Moderbacher CR, Gálvez RI, Cortes FH, Grifoni A, Tarke A, Chang J, Escarrega EA, Kim C, Goodwin B, Bloom NI, Frazier A, Weiskopf D, Sette A, Crotty S. Humoral and cellular immune memory to four COVID-19 vaccines. Cell. 2022 Jul 7;185(14):2434–2451.e17. doi: 10.1016/j.cell.2022.05.022.

