## Supplemental tables for "Safety and Immunogenicity of an Inactivated Recombinant Newcastle Disease Virus Vaccine Expressing SARS-CoV-2 Spike: A Randomised, Comparator-Controlled, Phase 2 Trial"

### Supplemental Table 1: Antibodies used in the AIM assay

| Reagent | Clone (Source) | Catalog No | Dilution |
|---|---|---|---|
| CD40 pure- functional grade | HB14 (Miltenyi Biotec) | 130-094-133 | 1.5:100 |
| CCR7 - BV711 | G043H7 (Biolegend) | 353228 | 1:200 |
| Fixable Live/Dead Blue | (Thermo) | L23105 | 1:1000 |
| Human FC Block | (BD) | 564220 | 1:20 |
| CD3 - BUV395 | UCHT1 (BD) | 563546 | 1:1000 |
| CD8 - BUV805 | SK1 (BD) | 612889 | 1:1000 |
| CD16 - BV510 | 3G8 (Biolegend) | 302048 | 1:1000 |
| CD14 - BV510 | 63D3 (Biolegend) | 367124 | 1:1000 |
| CD20 - BV510 | 2H7 (Biolegend) | 302340 | 1:1000 |
| CD45RA - BV570 | HI100 (Biolegend) | 304132 | 1:1000 |
| CD4 - cFluor b548 | SK3 (Cytek) | R7-20043 | 1:500 |
| PD-1 - BV785 | EH12.2H7 (Biolegend) | 329930 | 1:200 |
| CD69 - FITC | FN50 (Biolegend) | 310904 | 1:200 |
| CD40L - PE-Dazzle594 | 24-31 (Biolegend) | 310840 | 1:200 |
| CD137 - BUV737 | 4b4-1 (BD) | 741861 | 1:100 |
| OX40 - APC | Ber-Act35 (Biolegend) | 350008 | 1:100 |
| Brilliant Staining Buffer Plus | (BD) | 566385 | 1:10 |

### Supplement Table 2: Number of subjects with unsolicited adverse events with onset during 28 days after vaccination in the safety analysis population

| | COVIVAC | | |
|---|---|---|---|
| **Number of subjects with:** | **3 µg (N = 124)** n (%) (95% CI*) | **6 µg (N = 125)** n (%) (95% CI*) | **VAXZEVRIA (N = 125)** n (%) (95% CI*) |
| **One or more adverse events** | n= 124 | n= 125 | n= 125 |
| | 36 (29·0%) | 29 (23·2%) | 39 (31·2%) |
| | (21·2-37·9) | (16·1-31·6) | (23·2-40·1) |
| **Vaccine-related adverse events** | n= 124 | n= 125 | n= 125 |
| | 0 (0%) | 0 (0%) | 0 (0%) |
| | (0·0-2·9) | (0·0-2·9) | (0·0-2·9) |
| **Withdrawn due to an adverse event** | n= 124 | n= 125 | n= 125 |
| | 0 (0%) | 0 (0%) | 0 (0%) |
| | (0·0-2·9) | (0·0-2·9) | (0·0-2·9) |

**Note**: Two-sided 95% confidence intervals (CIs) computed via the Clopper-Pearson method.

**Supplement Table 3: Summary of geometric mean titer (GMT) of $NT_{50}$ against SARS-CoV-2 pseudovirus by age group in the full analysis population**

| $NT_{50}$ measure | | | COVIVAC 3 µg (N = 124) | COVIVAC 6 µg (N = 125) | VAXZEVRIA (N = 125) |
|---|---|---|---|---|---|
| Baseline (D1) | 18-59 yr. | GMT (95% CI) | n = 80 | n = 79 | n = 79 |
| | | | 5·04 (4·96 , 5·13) | 5·00 ( - ) | 5·23 (4·78 , 5·72) |
| | ≥ 60 yr. | GMT (95% CI) | n = 40 | n = 41 | n = 34 |
| | | | 5·00 ( - ) | 5·14 (4·86 , 5·45) | 5·12 (4·88 , 5·36) |
| 14 days after the second vaccination (D43) | 18-59 yr. | GMT (95% CI) | n = 80 | n = 78 | n = 79 |
| | | | 153·28 (124·22 , 189·15) | 176·52 (141·45 , 220·27) | 99·92 (80·80 , 123·56) |
| | ≥ 60 yr. | GMT (95% CI) | n = 40 | n = 40 | n = 35 |
| | | | 183·57 (133·40 , 252·61) | 257·87 (181·10 , 367·18) | 79·49 (55·68 , 113·46) |
| 6 months after the second vaccination (D197) | 18-59 yr. | GMT (95% CI) | n = 78 | n = 78 | n = 77 |
| | | | 188·05 (100·93 , 350·38) | 140·99 (76·57 , 259·61) | 126·74 (77·40 , 207·55) |
| | ≥ 60 yr. | GMT (95% CI) | n = 38 | n = 37 | n = 34 |
| | | | 98·94 (40·18 , 243·62) | 116·19 (47·80 , 282·45) | 50·00 (21·18 , 118·03) |

**Note**: - $NT_{50}$ GMTs are analyzed from subjects with seronegative anti-S IgG at baseline.

**Supplement Table 4: Percentage of subjects with $NT_{50}$ titer sero responses against SARS-CoV-2 pseudovirus as defined by a ≥ 4-fold increase from baseline by age group in the full analysis population**

| $NT_{50}$ measure ≥ 4-fold (Titer) | | | COVIVAC 3 µg (N = 124) | COVIVAC 6 µg (N = 125) | VAXZEVRIA (N = 125) |
|---|---|---|---|---|---|
| 14 days after the second vaccination (D43) | 18-59 yr. | n (%) (95% CI) | n = 82 | n = 83 | n = 82 |
| | | | 73 (89·0%) | 77 (92·8%) | 70 (85·4%) |
| | | | (80·2-94·9) | (84·9-97·3) | (75·8-92·2) |
| | ≥ 60 yr. | n (%) (95% CI) | n = 42 | n = 40 | n = 40 |
| | | | 41 (97·6%) | 38 (95·0%) | 30 (75·0%) |
| | | | (87·4-99·9) | (83·1-99·4) | (58·8-87·3) |
| 6 months after the second vaccination (D197) | 18-59 yr. | n (%) (95% CI) | n = 80 | n = 82 | n = 80 |
| | | | 44 (55·0%) | 39 (47·6%) | 44 (55·0%) |
| | | | (43·5-66·2) | (36·4-58·9) | (43·5-66·2) |
| | ≥ 60 yr. | n (%) (95% CI) | n = 40 | n = 37 | n = 37 |
| | | | 17 (42·5%) | 17 (45·9%) | 13 (35·1%) |
| | | | (27·0-59·1) | (29·5-63·1) | (20·2-52·5) |

**Note**: The 95% CIs are computed via the Clopper-Pearson method. The analysis included subjects regardless of anti S IgG status at baseline.

**Supplement Table 5: Summary of geometric mean titer ratio (GMT Ratio) of $NT_{50}$ against SARS-CoV-2 pseudovirus by age group in the full analysis population**

| $NT_{50}$ measure GMT ratio (95% CI) | | COVIVAC | | |
|---|---|---|---|---|
| | | 6 µg / 3 µg | 3 µg / VAXZEVRIA | 6 µg / VAXZEVRIA |
| Baseline (D1) | 18-59 yr. | 0·99 (0·97 , 1·01) | 0·96 (0·88 , 1·05) | 0·96 (0·88 , 1·04) |
| | ≥ 60 yr. | 1·03 (0·97 , 1·09) | 0·98 (0·94 , 1·02) | 1·01 (0·93 , 1·08) |
| 14 days after the second vaccination (D43) | 18-59 yr. | 1·15 (0·85 , 1·56) | 1·53 (1·14 , 2·06) | 1·77 (1·30 , 2·40) |
| | ≥ 60 yr. | 1·40 (0·88 , 2·24) | 2·31 (1·45 , 3·69) | 3·24 (1·98 , 5·32) |
| 6 months after the second vaccination (D197) | 18-59 yr. | 0·75 (0·32 , 1·78) | 1·48 (0·67 , 3·27) | 1·11 (0·51 , 2·43) |
| | ≥ 60 yr. | 1·17 (0·34 , 4·08) | 1·98 (0·58 , 6·77) | 2·32 (0·69 , 7·85) |

**Note**: $NT_{50}$ GMTs are analyzed from subjects with seronegative anti-S IgG at baseline.

**Supplement Table 6: GMT of $NT_{50}$ against vaccine-homologous SARS-CoV-2 pseudovirus (PNA) compared to live virus (LVNA) 14 days after the second vaccination (D43), with GMT ratio contrasting COVIVAC to AZ in the full analysis population**

| **$NT_{50}$ by PNA** | | | |
|---|---|---|---|
| **Statistic** | **COVIVAC 3** | **COVIVAC 6** | **VAXZEVRIA** |
| N | 120 | 118 | 114 |
| GMT | 162.78 | 200.72 | 93.14 |
| 95% CI | (136.79, 193.71) | (166.15, 242.47) | (77.70, 111.66) |
| **Statistic** | **COVIVAC 3/VAXZEVRIA** | **COVIVAC 6/VAXZEVRIA** | |
| GMT ratio | 1.75 | 2.15 | |
| 95% CI | (1.36, 2.24) | (1.66, 2.80) | |
| **$NT_{50}$ by LVNA** | | | |
| **Statistic** | **COVIVAC 3** | **COVIVAC 6** | **VAXZEVRIA** |
| N | 120 | 117 | 114 |
| GMT | 45.84 | 70.35 | 29.84 |
| 95% CI | (35.82, 58.66) | (55.91, 88.53) | (23.74, 37.52) |
| **Statistic** | **COVIVAC 3/VAXZEVRIA** | **COVIVAC 6/VAXZEVRIA** | |
| GMT ratio | 1.54 | 2.36 | |
| 95% CI | (1.10, 2.15) | (1.71, 3.26) | |

**Note**: $NT_{50}$ GMTs are analyzed from subjects with seronegative anti-S IgG at baseline.

**Supplement Table 7: Summary of geometric mean concentration (GMC) in BAU/mL of anti-S IgG assessed by ELISA by age group in the full analysis population**

| Anti-S IgG measure (BAU/mL) | | | COVIVAC | | VAXZEVRIA (N = 125) |
|---|---|---|---|---|---|
| | | | 3 µg (N = 124) | 6 µg (N = 125) | |
| Baseline (D1) | 18-59 yr. | GMC (95% CI) | n = 80 | n = 79 | n = 80 |
| | | | 3.15 ( - ) | 3·15 ( - ) | 3·15 ( - ) |
| | ≥ 60 yr. | GMC (95% CI) | n = 40 | n = 41 | n = 35 |
| | | | 3·15 ( - ) | 3·15 ( - ) | 3·15 ( - ) |
| 14 days after the second vaccination (D43) | 18-59 yr. | GMC (95% CI) | n = 80 | n = 78 | n = 79 |
| | | | 139·60 (112·48 , 173·27) | 155·46 (123·39 , 195·87) | 366·41 (306·59 , 437·91) |
| | ≥ 60 yr. | GMC (95% CI) | n = 40 | n = 40 | n = 35 |
| | | | 146·65 (104·62 , 205·56) | 224·91 (155·86 , 324·57) | 312·37 (211·23 , 461·93) |
| 6 months after the second vaccination (D197) | 18-59 yr. | GMC (95% CI) | n = 78 | n = 78 | n = 77 |
| | | | 166·99 (87·31 , 319·39) | 124·64 (65·78 , 236·19) | 277·29 (164·97 , 466·07) |
| | ≥ 60 yr. | GMC (95% CI) | n = 40 | n = 39 | n = 34 |
| | | | 78·28 (30·39 , 201·61) | 89·87 (35·08 , 230·22) | 107·22 (42·89 , 268·03) |

**Note**: GMCs are analyzed from subjects with seronegative anti-S IgG at baseline.

**Supplement Table 8: Summary of geometric mean concentration ratio (GMC Ratio) of anti-S IgG assessed by ELISA by age group in the full analysis population**

| Anti-S IgG GMC ratio (95% CI) | | COVIVAC | | |
|---|---|---|---|---|
| | | **6 µg / 3 µg** | **3 µg / VAXZEVRIA** | **6 µg / VAXZEVRIA** |
| Baseline (D1) | 18-59 yr. | 1·00 (1·00 , 1·00) | 1·00 (1·00 , 1·00) | 1·00 (1·00 , 1·00) |
| | ≥ 60 yr. | 1·00 (1·00 , 1·00) | 1·00 (1·00 , 1·00) | 1·00 (1·00 , 1·00) |
| 14 days after the second vaccination (D43) | 18-59 yr. | 1·11 (0·81 , 1·52) | 0·38 (0·29 , 0·50) | 0·42 (0·32 , 0·57) |
| | ≥ 60 yr. | 1·53 (0·94 , 2·51) | 0·47 (0·28 , 0·78) | 0·72 (0·42 , 1·22) |
| 6 months after the second vaccination (D197) | 18-59 yr. | 0·75 (0·30 , 1·84) | 0·60 (0·26 , 1·37) | 0·45 (0·20 , 1·02) |
| | ≥ 60 yr. | 1·15 (0·31 , 4·27) | 0·73 (0·20 , 2·70) | 0·84 (0·23 , 3·07) |

**Note**: Anti-S IgG GMCs ratio are analyzed from subjects with seronegative anti-S IgG at baseline.

**Supplement Table 9: Summary of geometric mean fold rise (GMFR) from baseline of anti-S IgG concentration assessed by ELISA by age group in the full analysis population**

| Anti-S IgG measure | | | COVIVAC 3 µg (N = 124) | COVIVAC 6 µg (N = 125) | VAXZEVRIA (N = 125) |
|---|---|---|---|---|---|
| 14 days after the second vaccination (D43) | 18-59 yr. | GMFR from baseline (95% CI) | n = 81 | n = 82 | n = 83 |
| | | | 45·18 (36·37 , 56·12) | 46·55 (37·08 , 58·43) | 108·47 (89·94 , 130·83) |
| | ≥ 60 yr. | GMFR from baseline (95% CI) | n = 42 | n = 41 | n = 41 |
| | | | 45·03 (32·46 , 62·45) | 69·32 (48·25 , 99·59) | 87·13 (60·32 , 125·84) |
| 6 months after the second vaccination (D197) | 18-59 yr. | GMFR from baseline (95% CI) | n = 79 | n = 83 | n = 81 |
| | | | 54·93 (28·85 , 104·60) | 39·87 (21·68 , 73·30) | 78·74 (47·13 , 131·54) |
| | ≥ 60 yr. | GMFR from baseline (95% CI) | n = 42 | n = 39 | n = 40 |
| | | | 22·64 (9·12 , 56·18) | 28·52 (11·13 , 73·06) | 35·38 (15·15 , 82·63) |

**Note**: Geometric Mean Fold Rise (GMFR) is the geometric mean of the ratios of post-vaccination to the pre-first vaccination at D1 (baseline).
